# First imported Cases of MPXV Clade Ib in Goma, Democratic Republic of the Congo: Implications for Global Surveillance and Transmission Dynamics

**DOI:** 10.1101/2024.09.12.24313188

**Authors:** Daniel Mukadi-Bamuleka, Eddy Kinganda-Lusamaki, Noella Mulopo-Mukanya, Adrienne Amuri-Aziza, Áine O’Toole, Brigitte Modadra-Madakpa, Guy Mutombo Ndongala, Emmanuel Hasivirwe Vakaniaki, Sydney Merritt, Cris Kacita, Gaston Lubambo Maboko, Jean-Claude Makangara-Cigolo, Michel Ngimba, Emmanuel Lokilo, Elisabeth Pukuta-Simbu, Gradi Luakanda, Tavia Bodisa-Matamu, Zephanie Paluku Kalimuli, Prince Akil-Bandali, Sifa Kavira, Daan Jansen, Adèle Kavira Kamaliro, Emile Muhindo-Milonde, Jeriel Mufungizi, Yves Birindwa Hamisi, Hugo Kavunga, Olivier Tshiani, Sabin S. Nundu, Laurens Liesenborghs, Nicole A. Hoff, Jean Nachega, Robert Shongo, Ahidjo Ayouba, Genay Pilarowski, Alain Kakule Mangolopa, Amos Kiuka Ebondo, Nicola Low, Souradet Y. Shaw, Sam Wilkinson, Sofonias Kifle Tessema, Lorenzo Subissi, Eric Delaporte, Koen Vercauteren, Tony Wawina-Bokalanga, Anne W Rimoin, Martine Peeters, Nicholas Loman, Andrew Rambaut, Jean-Jacques Muyembe-Tamfum, Lisa E. Hensley, Jason Kindrachuk, Placide Mbala-Kingebeni, Steve Ahuka-Mundeke

## Abstract

The ongoing national mpox outbreak in the Democratic Republic of the Congo has resulted in more >30,000 suspected cases in the country from January 2023 to August 2024. While these historic case totals have been driven by primarily by zoonosis, the emergence of Clade Ib monkeypox virus (MPXV), which is connected to more sustained human-to-human transmission, has been associated with increasing public health impacts in eastern DRC. First identified in South Kivu province, Clade Ib MPXV has been identified in multiple non-endemic East African countries for the first time. In DRC, there have been concerns over broader Clade Ib expansion in the country that could further complicate containment and mitigation responses. Here, we report the first introductions of Clade Ib into North Kivu province, including within internal displacement camps, with suspected close contact transmission that includes non-intimate contacts and children. These findings demonstrate that mpox case investigations and community messaging campaigns should include considerations for non-sexual contact-mediated transmission of Clade Ib that includes children <15 years.

## INTRODUCTION

Mpox, an emerging zoonotic disease caused by Monkeypox virus (MPXV), garnered significant international attention in 2022 due to virus expansion and broad human-to-human transmission concentrated within dense sexual networks in more than 100 historically non-endemic countries (1), which resulted in a first declaration of a public health emergency of international concern (PHEIC) by the World Health Organization (WHO) in the same year (2–4). Human mpox was first described in 1970 in the Democratic Republic of the Congo (DRC) and is endemic among tropical forested regions of Central and West Africa (5). While outbreaks have been sporadic historically, there has been a generally increasing burden of disease across endemic areas with the DRC facing the greatest public health impact (2, 6, 7). This has coincided with decreasing immunity to human orthopoxviruses among the population over time following the cessation of the global smallpox vaccination program, and in increasing immune naive population (8–10). In 2024, a surge in cases in the African region which followed the emergence of a new MPXV variant resulted in a second declaration of a PHEIC (11).

Historically, zoonosis has been the primary driver of human mpox within endemic regions with rodent species being the presumed reservoir and limited secondary transmission among close contacts. However, sustained human-to-human transmission has been increasingly associated with MPXV infections such as in Nigeria following re-emergence of the virus in 2017 and during the global 2022 mpox outbreak (12, 13). Close sexual (intimate) contact and altered or atypical clinical disease presentation was highly overrepresented among cases during the global outbreak (3, 14), with 96% of cases being males and 87% of cases globally self-identifying as MSM (15).

Since early 2023, there has been an ongoing historic national mpox outbreak in DRC that has included virus expansion and transmission within communities in adjacent non-endemic countries. Notably, MPXV infections associated with sustained human-to-human transmission including sexual (intimate) contact have been reported for the first time in Kwango and South Kivu provinces within the country (16, 17). In addition, suspected mpox cases have been reported in 25 of 26 of the provinces in DRC including multiple large urban centers. This has also included sustained virus transmission chains within regions having infrequent reports of suspected mpox cases. Notably, mpox has rapidly expanded in South Kivu province, with cases increasing from 10 suspected cases per week to >381 suspected cases per week in epidemiological week 31, 2024 (18). In Kamituga Health Zone, a mining region in South Kivu province, mpox cases were first reported in September 2023, with 51.9% of cases identified in women, 29% among professional sex workers (PSW), and a median age of 22 years among confirmed cases (16). We identified APOBEC3-like mutations in high-quality complete MPXV genomes from Kamituga, which led us to recommend the subdivision of Clade I MPXV into subclades Ia and Ib, with the latter related to sustained human-to-human transmission trends (16), and the former predominantly linked to zoonotic transmission (19). Estimates from molecular clock analysis suggest that Clade Ib MPXV has been circulating locally in Kamituga since mid-September 2023 (95% highest posterior density intervals July 2023-October 2023) (16).

Given the ongoing historic impacts of mpox in DRC and further geographic expansion, the increasing burden of Clade Ib-associated infections, ongoing internal displacement due to armed conflicts, and the commercial activities within the region, there is a critical need for expanded mpox surveillance in Eastern DRC (20). Here, we describe the observed expansion of Clade Ib MPXV from South Kivu into North Kivu Province, including mpox circulation within internal displacement camps. Our results also provide evidence for Clade Ib transmission associated with both sexual (intimate) and non-sexual close contacts, including among children, with important implications for infection prevention and control recommendations in the recent declaration of the second PHEIC for mpox.

## MATERIALS AND METHODS

### Ethical Considerations

This study was exempted from ethical approval since it was conducted as part of the national surveillance activities and carried out in the public interest by the Ministry of Health, the Democratic Republic of the Congo. All activities described were undertaken as part of regular public health surveillance conducted and approved directly by the Ministry of Health, Democratic Republic of the Congo. Data was provided by the National Programme for Control of Mpox and Viral Haemorrhagic Fevers and the National Institute for Biomedical Research, the Democratic Republic of the Congo, as part of the case investigations. No written informed consent for research was provided as the analyses conducted in this study were done retrospectively and the information and diagnostic samples were collected for surveillance and clinical care purposes.

### Suspected mpox case investigations

Human mpox is a mandatory reportable disease in the DRC, with an established case definition from the Ministry of Health which has been in use since 2001. This definition was expanded upon in 2010 to better enhance surveillance in Tshuapa Province. In this expanded case definition, a suspected case was defined as follows: a patient with a vesicular pustular eruption characterized by hard and deep pustules and with at least one of the following symptoms: fever preceding the eruption, lymphadenopathy (inguinal, axillary, or cervical), and/or pustules or crusts on the palms of the hands or soles of the feet, and having an exposure such as a travelling history from an affected area, a high risk contact with people coming from affected area or exposure to wild animal dead or with lesions (21). Formal investigation of a suspected mpox case included the collection of samples and the completion of a case investigation form by trained staff from Health zone and Provincial Health division (DPS).

### Sample collection and laboratory diagnosis of mpox

Lesion swabs and/or lesion crusts are the preferred samples for MPXV diagnostics, but blood samples and nasopharyngeal swabs were also collected. These samples were then shipped to the Rodolphe-Merieux INRB-Goma Laboratory for PCR assay and whole genome sequencing (WGS). To confirm the diagnosis of mpox, the GeneXpert platform was used with clade IIb-specific cartridges, according to the manufacturer’s instructions. Mpox-positive samples with a Ct value < 30 were selected for subsequent WGS.

### Bioinformatics analysis

FASTQ files from GridION were base called with the High accuracy model from Guppy v6, and reads were demultiplexed and adapter-trimmed by the GridION built-in MinKNOW software. MPXV consensus genomes were generated using the artic (https://github.com/artic-network/artic-mpxv-nf) and metatropics pipelines (https://github.com/DaanJansen94/nextflow-metatropics-INRB).

### Phylogenetic and APOBEC-3 analysis

We estimated a maximum likelihood phylogeny using IQ-TREE 2 version 2.2.5 (22) with the Hasegawa, Kishino, Yano (HKY) substitution model. Ancestral reconstruction was performed for each internal node on phylogeny using IQ-TREE 2, enabling mapping of single nucleotide polymorphisms (SNPs) along branches. SNPs were categorized on the basis of whether they were consistent with the signature of APOBEC3 editing, assuming this process induced specific mutations (TC → TT and GA → AA) as previously described (23, 24).

## RESULTS

### Case investigation and epidemiological assessment

We describe a total of nine confirmed mpox cases identified in North Kivu, DRC, which includes six cases confirmed as Clade Ib MPXV by whole genome sequencing. A map of the geographic locations of the identified cases is presented in Figure 1. Epidemiological links among Cases 1-9 are presented in Figure 2.

**Figure 1:**
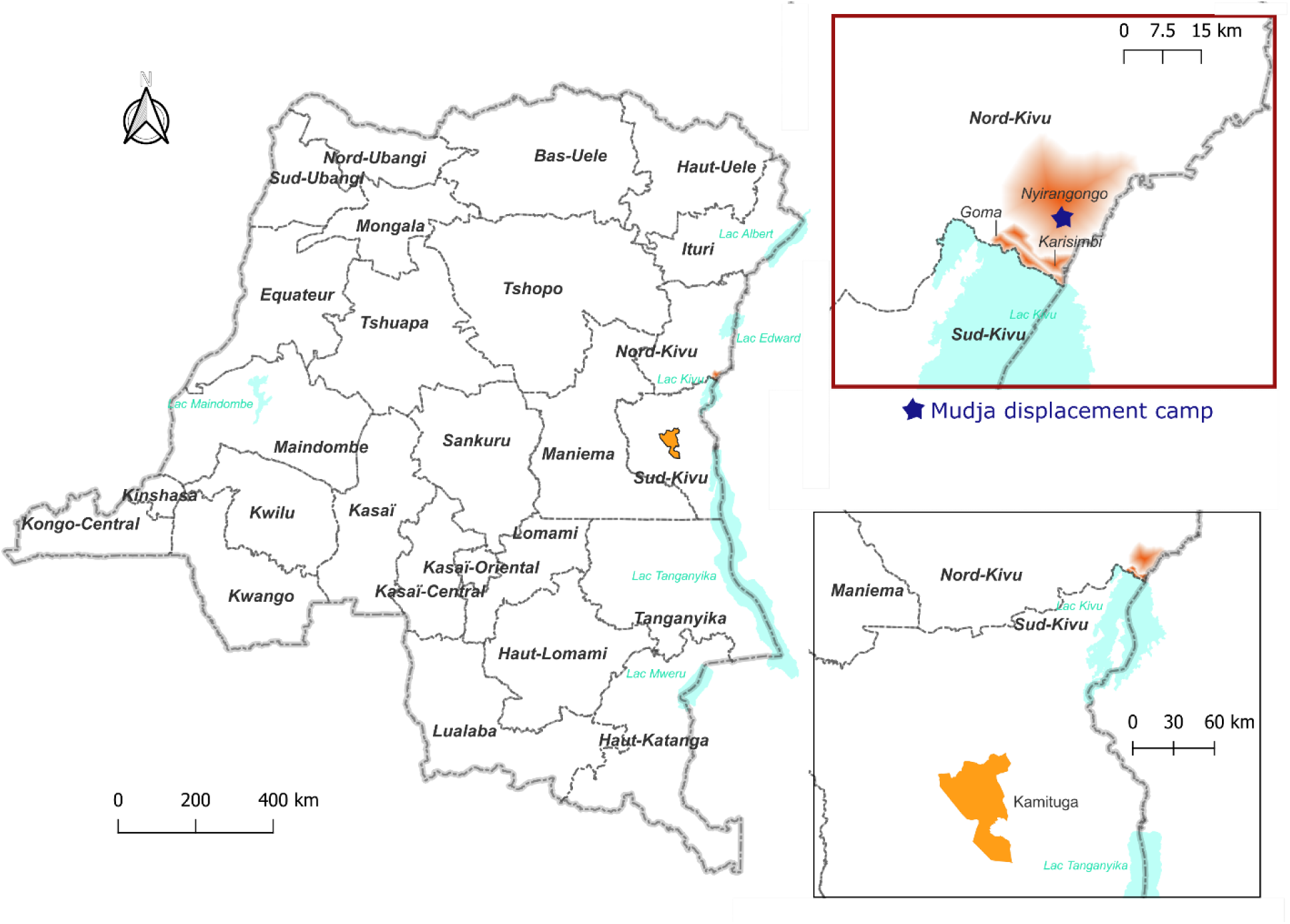
Geographic locations of cases identified in North Kivu including Mudja displacement camp. Map constructed using QGIS3.22.11. The fading red coloring on the inset signifies the three Goma health zones where the sequenced samples originated from. The star representing Mudja (or Muja) camp location within Nyiragongo health zone utilizing coordinates from Google maps and OpenStreetMap.

**Figure 2:**
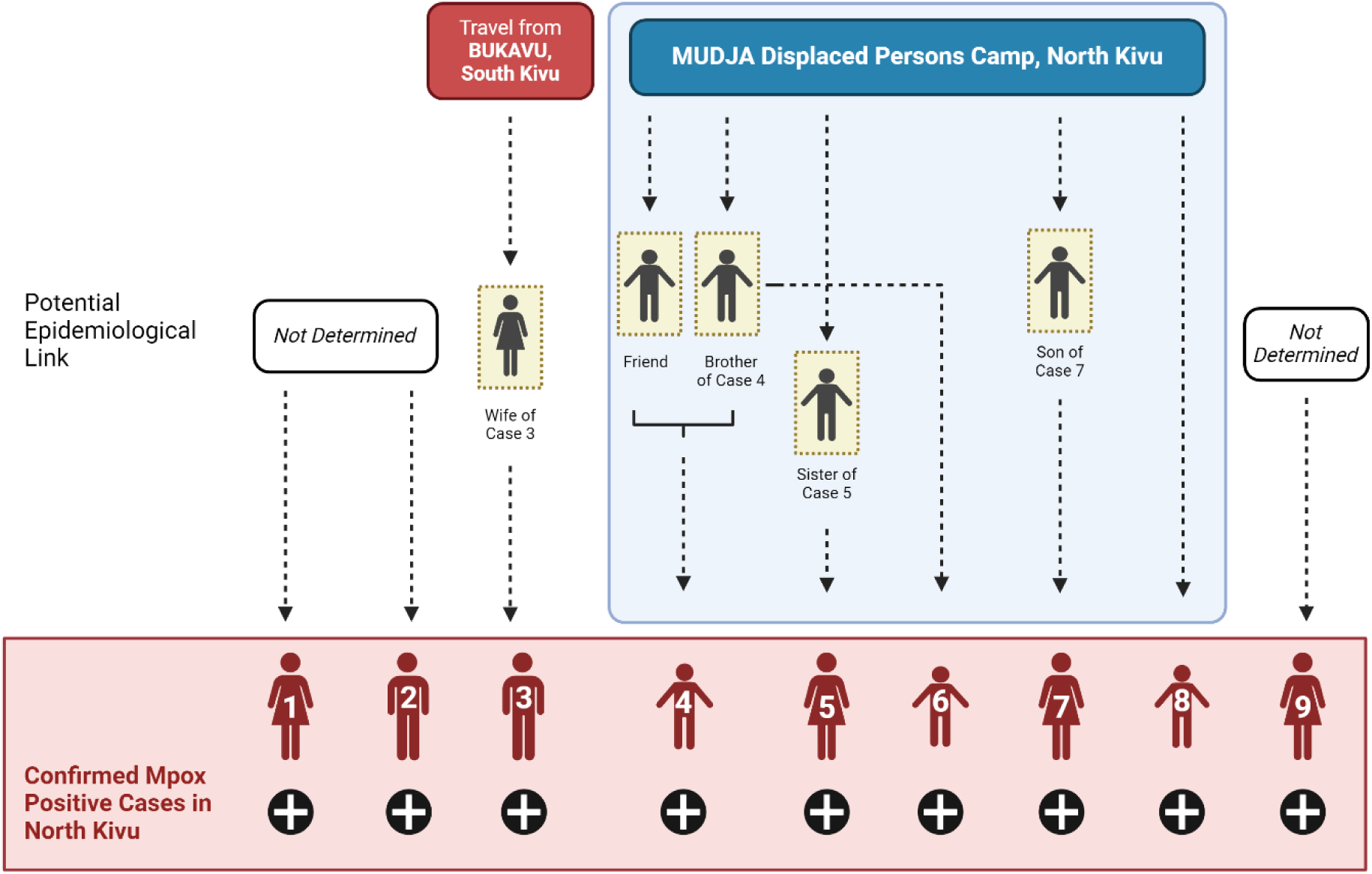
Epidemiological linkages and suspected transmission chains among Clade Ib mpox cases identified in Goma, North Kivu, DRC. Transmission chain at the Division Provincial de la Santé (DPS), North Kivu. Clade Ib MPXV has been confirmed for cases 1-3 and results are pending for cases 4-9 at this time.

Case 1, an adult female (15-30 years) from North Kivu, developed vesicular lesions and pustules on day 5 post-symptom onset and healthcare consultation in May 2024. The patient was discharged one day later (day 6) and cared for at home. Vesicle and nasopharyngeal swabs were collected on day 8 post-symptom onset and confirmed positive for orthopoxvirus by PCR. Additional tests for syphilis rapid plasma reagin (RPR) and HIV (determine) were negative. The individual reported travel to South Kivu, which is currently affected by an ongoing Clade Ib MPXV outbreak, during the 21 days prior to symptom development. However, this could not be confirmed. Follow up investigations of contacts from the healthcare consultation (n=23) and at their residence (n=19) did not identify any additional mpox cases.

Case 2, an adult male, 15-30 years from North Kivu, received consultation for management of vesicular lesions, which were in the process of healing in June 2024. Lesion swabs were collected and confirmed positive for mpox one day following consultation. Sexual intercourse within 21 days of symptom onset with an occasional sexual partner from North Kivu was disclosed during case investigation though the individual did not notice any visible lesions on the partners’ body.

Case 3, an adult male aged 15-30 years from North Kivu received medical consultation eight days post-symptom onset. Mpox was confirmed by PCR one day following consultation. The individual reported that his spouse had received consultation for similar symptoms. Through this, the case investigation established an epidemiological link with his spouse, a probable mpox case, that included recent travel to South Kivu in the prior 21 days (probable case 1). There were no epidemiological links established between Cases 1-3.

Five additional confirmed mpox cases, Cases 4-8, were residents of Mudja displaced persons camp, North Kivu. Probable epidemiological links were made among Cases 4-6 based on frequent interactions among residents within the camp, including probable cases.

Case 4, a male child <15 years, developed fever and headache. The child reported common use of a sponge that had been used on his male sibling (<15 years) who recently had dermatological lesions similar to Case 4 though no confirmatory testing had been performed (probable case 2). The male sibling of Case 4 had also interacted with a neighbor who had developed skin lesions a few days prior to the sibling though no further information or testing information was available (probable case 3). It was also noted that Case 4 and their male sibling had shared sleeping quarters. Skin eruptions including vesicles and pustules were noted for Case 4 on day 3 post-symptom onset; mpox was confirmed from samples taken on day 5.

Case 5, a female 15-30 years, developed symptoms in mid-June 2024 including chills, cough, headache, lymphadenopathy, eruptions on the lips, and pubic lesions. A female child living within the same household had reportedly developed similar symptoms ∼14 days earlier but had not sought clinical consultation (probable case 4).

Case 6, a male child <15 years, presented with lesions on the back, face and abdomen. Case 6 was friends with probable case 2.

Case 7, a female >30 years, developed a rash on the buttock, thigh, hands, and feet at symptom onset. Case 7 had no epidemiological link to other cases in Mudja camp; however, she reported having provided recent care for her son (<15 years;) who had a similar illness presentation and with whom she shared a bed (probable case 5). Probable epidemiological links were made among Cases 4-6 based on frequent interactions among residents within the camp, including probable cases.

Case 8, a female child (<15) living in Mudja camp, had fever at symptom onset followed by vesicular eruptions on the cuisses, abdomen, back and face, with no adenopathy. An epidemiological link has not been clearly established with other probable or confirmed cases in the camp; however, they had contact with other children through regular plays activities.

Case 9, a 15-30-year-old female, presented with vesiculo-pustular lesions on day 3 post-symptom onset (fever starting on day 0). Samples were taken for testing on day 10 and confirmed positive for MPXV. There were no epidemiological links to any of the other cases, confirmed or probable.

### Case demographics and clinical symptoms

Demographic and clinical data for all nine confirmed mpox cases are presented in Table 1. Of all cases, five (5/9) were residents at the Mudja displaced persons site for internally displaced people. Males comprised 5/9 cases; 4/9 cases were female. The mean age was 18 years (6 - 45 years). The majority of cases were identified among those aged 15-30 years (5/9) and three cases were identified among those <15 years. Most patients (6/9) required hospitalization and included three males and three females. Of those hospitalized, 3/6 were aged 15-30 years, 2/6 were <15 years, and 1/6 was >30 years. There were no fatal infections recorded among the nine confirmed cases.

**Table 1:** Demographic and clinical symptom data for mpox cases identified in Goma to date. Column percentages are presented in parentheses.

| *Age group (years)* | **Male** | **Female** | **Total** |
| --- | --- | --- | --- |
| **<15** | 2 | 1 | 3 |
| **15-30** | 2 | 3 | 5 |
| **>30** | 1 | 0 | 1 |
| **Total** | **5** | **4** | **9** |
| Hospitalization | 3 (60%) | 3 (75%) | 6 (67%) |
| *Clinical symptoms* |  |  |  |
| **Cutaneous eruptions** | 5 (100%) | 4 (100%) | 9 (100%) |
| **Genital eruptions** | 4 (80%) | 3 (75%) | 7 (78%) |
| **Oral eruptions** | 2 (40%) | 4 (100%) | 6 (67%) |
| **Fever** | 3 (60%) | 3 (75%) | 6 (67%) |
| **Headache** | 4 (80%) | 3 (75%) | 7 (78%) |
| **Myalgia** | 5 (100%) | 3 (75%) | 8 (89%) |
| **Arthralgia** | 4 (80%) | 3 (75%) | 7 (78%) |
| **Fatigue** | 1 (20%) | 2 (50%) | 3 (33%) |
| **Cervical lymphadenopathy** | 3 (60%) | 4 (100%) | 7 (78%) |
| **Inguinal lymphadenopathy** | 3 (60%) | 2 (50%) | 5 (56%) |
| **Axillary lymphadenopathy** | 2 (40%) | 1 (25%) | 3 (33%) |

Cutaneous eruptions were reported for all patients (9/9) with genital and oral eruptions frequently reported (7/9 and 6/9 cases, respectively). Genital eruptions were similarly reported whether male or female (4/5 and 3/4 cases, respectively); however, oral eruptions were reported more frequently among female cases than males (4/4 and 2/4 cases, respectively). Myalgia (8/9), headache, (7/9), and fever (6/9) were common among mpox patients. Lymphadenopathy was also frequently reported with cervical lymphadenopathy being the most commonly reported among cases (7/9) as compared to inguinal (5/9) or axillary (3/9).

### Genome sequencing identifies Clade Ib introduction into Goma, North Kivu

Genomic analysis of the first three confirmed cases showed that they all clustered within MPXV Clade Ib together with Mpox cases detected in South Kivu (Figure 3). Their position in the tree with MPXV sequences from South Kivu suggests they were part of the sustained human outbreak first reported in Kamituga health zone. These findings are consistent with the reported travel of the first case from South Kivu. The genomes from the cases 1 & 2 are closely linked suggesting they are part of the same transmission chain, although no epidemiological link between the two was proven by the investigation. The third sequenced case is separated from the first two in the Clade Ib outbreak tree, implying an independent introduction into Goma.

**Figure 3:**
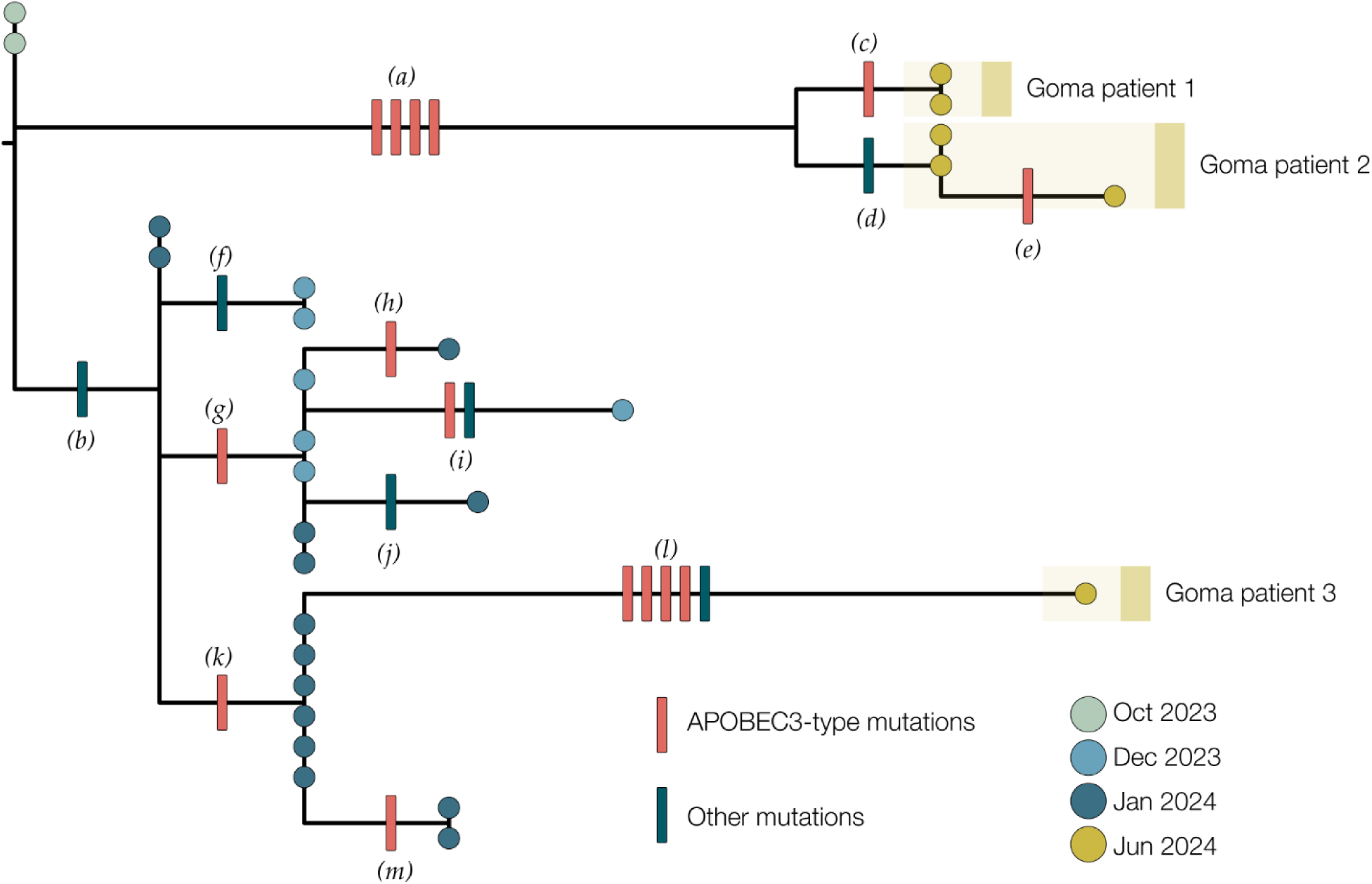
A maximum likelihood tree constructed using IQ-Tree 2 (22) with the HKY substitution model (26). We included a Clade Ia genome as an outgroup and then removed it after rooting. Single nucleotide mutations are reconstructed and displayed denoting whether they are APOBEC3-mediated (red bars) or other mutations (blue bars). Genomes from recent cases in Goma, North Kivu are denoted with yellow circles. Patients 1 and 2 each have multiple genomes sequenced from different samples. Blue circles are genomes from Kamituga, South Kivu from October and December 2023, and January 2024. Letters in parentheses are referenced in Table 2.

**Table 2.** List of mutations in the 2023-2024 DRC mpox outbreak.

| **Branch label<sup>1</sup>** | **Genome position<sup>2</sup>** | **Mutation<sup>3</sup>** | **APOBEC3?** |
| --- | --- | --- | --- |
| (a) | 103,660 | GA->AA | yes |
|  | 130,155 | GA->AA | yes |
|  | 142,587 | GA->AA | yes |
|  | 161,978 | TC->TT | yes |
| (b) | 165,597 | C->A | no |
| *(c)* | 64,117 | GA->AA | yes |
| *(d)* | 30,660 | T->C | no |
| *(e)* | 181,268 | GA->AA | yes |
| *(f)* | 167,805 | A->G | no |
| *(g)* | 115,484 | GA->AA | yes |
| *(h)* | 57,332 | GA->AA | yes |
| *(i)* | 89,000 | GA->AA | yes |
|  | 127,619 | A->G | no |
| *(k)* | 183,181 | G->A | no |
| *(l)* | 71,751 | TC->TT | yes |
|  | 139,485 | TC->TT | yes |
|  | 148,221 | GA->AA | yes |
|  | 168,749 | GA->AA | yes |
|  | 169,501 | T->A | no |

<sup>1</sup> Branch labels refer to Figure 3
<sup>2</sup> Coordinates relative to Clade I reference genome, NCBI accession NC\_003310
<sup>3</sup> Dinucleotide mutations are ascribed to APOBEC3 mutations

Of the 21 single nucleotide mutations reconstructed in this tree (Table 2), 15 are of the type expected due to the action of human APOBEC3. The viruses sequenced from cases 1 & 2 are descendants of viruses present in Kamituga in October 2023 or earlier. Furthermore, the branch leading to cases 1 & 2 has four APOBEC3-type mutations which even at the elevated rate of evolution induced by APOBEC3 would represent months of human transmission. In combination with the reported recent travel history, this indicates a likely recent introduction into the Goma area from the South Kivu region with ongoing transmission.

The branch leading to case 3 has four APOBEC3 mutations and one other mutation, possibly the result of an error during replication, indicating a similar timespan for this branch. However, this branch joins the Kamituga tree clustering with viruses sampled in January 2024, suggesting a direct link to that outbreak. The long unbroken branch with 5 single nucleotide mutations, and the fact that this lineage has not yet been sampled in Kamituga, may either be explained by undersampling in Kamituga, or an unresolved epidemiological link to South Kivu (i.e. by not having identified nor sequenced the contact of Case 3 who may also be epidemiologically linked to South Kivu).

However, the third hypothesis, being that the virus has been circulating in Goma for weeks or months, can also not be entirely excluded.

## DISCUSSION

Here, we present the first case series of Clade Ib mpox reported in North Kivu province, DRC, with probable introduction to the Mudja internal displacement camp. Case investigations and epidemiological analysis demonstrates non-sexual contact mediated transmission of Clade Ib MPXV that included cases among children. The investigations also suggested potential linkage of one of the mpox cases to recent travel to South Kivu, which has recently reported mpox cases. The introduction and expansion of mpox in large urban centers in DRC including Bukavu (urban pop. >1 million) and Goma (urban pop. ∼2 million) further increase the risk for greater public health impacts from the ongoing outbreak, which has already far surpassed the scale of prior outbreaks across endemic regions. Of particular note is the identification of MPXV Clade Ib circulation, which is linked to sustained human-to-human transmission chains, among both urban centers (16, 25). This risk is further compounded by Goma’s role as a major commercial hub and regional and international connection point, including air travel. Additionally, Goma’s proximity to international borders with East African countries and frequent unmonitored cross-border movement increases the potential for the virus to spread to new regions through cryptic transmission.

A concerning finding from this investigation was the identification of mpox cases among individuals at the Mudja displaced persons site. The introduction of MPXV to, and circulation within, displaced persons sites could have deleterious public health impacts. Importantly, this includes the potential for broad disease transmission given the poor sanitation conditions, highly dense populations, and very limited healthcare or surveillance support. Thus, ongoing political instability in the region could further destabilize mpox containment and mitigation efforts. Case investigations among the nine confirmed mpox cases in this study identified potential contacts with unreported symptomatic infections within households, highlighting the need for greater community engagement focused on both case recognition and suspected case reporting. Additionally, the case investigations also noted the potential for contacts among children through common play areas. Given the cases identified herein among children, and considering the disproportionate disease severity associated with mpox among children, additional vigilance should be undertaken to inform communities regarding the potential risks for mpox within children. Of important note was the potential linkage of Case 1 to recent travel to Bukavu, South Kivu, where there have been increasing detrimental public health impacts from mpox linked to sexual (intimate) contacts. Ongoing conflict within the region could also impact containment and mitigation efforts for MPXV circulation within Mudja given the potential for onward transmission among current residents as well as further introductions of virus through undiagnosed infections. Clade Ib introduction into Mudja is currently being assessed by viral genome sequencing from samples collected during this investigation.

Our investigation also demonstrated that infection risks for the recently identified MPXV Clade Ib extend beyond sexual (intimate) contacts to include caregivers and children as well as adults. Taken together, these factors create a critical bottleneck for response to public health emergencies reminiscent of those encountered in the region during the Ebola virus disease outbreak in 2018-2020. While the DRC already faces expansive public health hurdles, including extreme economic and development hardships, these impacts are further elevated within internal displacement sites (camps). The convergence of resource limitations within these sites including healthcare access, sanitation, clean water, and food as well as overcrowding will likely further facilitate rapid mpox circulation and broaden this outbreak to a larger humanitarian crisis. Consequently, the high risk of the mpox outbreak further expanding nationally and internationally must be considered an urgent issue and a priority for all stakeholders. These cases involved adult males and females, as well as children, who were infected by MPXV through various transmission routes, including close non-sexual contacts. Our analysis demonstrates the further expansion of Clade Ib MPXV and the first identification of cases within a displaced persons’ site highlighting concerns for rapid expansion of the outbreak among highly vulnerable populations.

The spread of Clade Ib MPXV in the city of Goma, North Kivu, is highly concerning, as the virus has the potential to expand further geographically within the DRC as well as more broadly internationally. This risk is exacerbated due to Goma’s proximity to porous international border regions, massive population displacement due to armed conflicts, and its international airport. Therefore, there is an urgent need for collaborative efforts and actions to combat mpox before its further spread to other countries.

## Data Availability

All data produced in the present study are available upon reasonable request to the authors

## ACKNOWLEDGEMENTS

This work was funded by the Department of Defense, Defense Threat Reduction Agency, Monkeypox Threat Reduction Network; and USDA Non-Assistance Cooperative Agreement #20230048; Africa Pathogen Genomics Initiative helped acquiring and maintaining the sequencer; Agence Française de Dévelopement through the AFROSCREEN project (grant agreement CZZ3209, coordinated by ANRS-MIE Maladies infectieuses émergentes in partnership with Institut de Recherche pour le Développement (IRD) and Pasteur Institute) for laboratory support and PANAFPOX project funded by ANRS-MIE ; Belgian Directorate-general Development Cooperation and Humanitarian Aid and the Research Foundation - Flanders (FWO, grant number G096222 N to L.L.); the International Mpox Research Consortium (IMReC) through funding from the Canadian Institutes of Health Research and International Development Research Centre (grant no. MRR-184813); US NIAID/NIH grant number U01AI151799 through Center for Research in Emerging Infectious Disease-East and Central Africa (CREID-ECA); E.L. received a PhD grant from the French Foreign Office. We acknowledge the support of the Wellcome Trust (Collaborators Award 206298/Z/17/Z, ARTIC network).

## REFERENCES

1. El Eid R, Allaw F, Haddad SF, Kanj SS. Human monkeypox: A review of the literature. PLoS Pathog. 2022;18(9):e1010768.

2. Van Dijck C, Hoff NA, Mbala-Kingebeni P, Low N, Cevik M, Rimoin AW, et al. Emergence of mpox in the post-smallpox era-a narrative review on mpox epidemiology. Clin Microbiol Infect. 2023;29(12):1487–92.

3. Thornhill JP, Gandhi M, Orkin C. Mpox: The Reemergence of an Old Disease and Inequities. Annu Rev Med. 2024;75:159–75.

4. Dodd LE, Lane HC, Muyembe-Tamfum JJ. Vaccines for Mpox - An Unmet Global Need. NEJM Evid. 2024;3(3):EVIDe2300348.

5. Marennikova SS, Seluhina EM, Mal’ceva NN, Cimiskjan KL, Macevic GR. Isolation and properties of the causal agent of a new variola-like disease (monkeypox) in man. Bull World Health Organ. 1972;46(5):599–611.

6. Bunge EM, Hoet B, Chen L, Lienert F, Weidenthaler H, Baer LR, et al. The changing epidemiology of human monkeypox-A potential threat? A systematic review. PLoS Negl Trop Dis. 2022;16(2):e0010141.

7. Simpson K, Heymann D, Brown CS, Edmunds WJ, Elsgaard J, Fine P, et al. Human monkeypox - After 40 years, an unintended consequence of smallpox eradication. Vaccine. 2020;38(33):5077–81.

8. Rimoin AW, Mulembakani PM, Johnston SC, Lloyd Smith JO, Kisalu NK, Kinkela TL, et al. Major increase in human monkeypox incidence 30 years after smallpox vaccination campaigns cease in the Democratic Republic of Congo. Proc Natl Acad Sci U S A. 2010;107(37):16262–7.

9. Simpson K, Heymann D, Brown CS, Edmunds WJ, Elsgaard J, Fine P, et al. Human monkeypox - After 40 years, an unintended consequence of smallpox eradication. Vaccine. 2020;38(33):5077–81.

10. Shah HH, Molani MK, Shabbir N. Human monkeypox - After 40 years, an unintended consequence of smallpox eradication. Front Public Health. 2022;10:1082586.

11. WHO Director-General declares mpox outbreak a public health emergency of international concern [press release]. 14 August 2024 2024.

12. Ndodo N, Ashcroft J, Lewandowski K, Yinka-Ogunleye A, Chukwu C, Ahmad A, et al. Distinct monkeypox virus lineages co-circulating in humans before 2022. Nat Med. 2023;29(9):2317–24.

13. Yinka-Ogunleye A, Aruna O, Dalhat M, Ogoina D, McCollum A, Disu Y, et al. Outbreak of human monkeypox in Nigeria in 2017-18: a clinical and epidemiological report. Lancet Infect Dis. 2019;19(8):872–9.

14. McCollum AM, Damon IK. Human monkeypox. Clin Infect Dis. 2014;58(2):260–7.

15. Laurenson-Schafer H, Sklenovska N, Hoxha A, Kerr SM, Ndumbi P, Fitzner J, et al. Description of the first global outbreak of mpox: an analysis of global surveillance data. Lancet Glob Health. 2023;11(7):e1012–e23.

16. Vakaniaki EH, Kacita C, Kinganda-Lusamaki E, O’Toole A, Wawina-Bokalanga T, Mukadi-Bamuleka D, et al. Sustained human outbreak of a new MPXV clade I lineage in eastern Democratic Republic of the Congo. Nat Med. 2024.

17. Kibungu EM, Vakaniaki EH, Kinganda-Lusamaki E, Kalonji-Mukendi T, Pukuta E, Hoff NA, et al. Clade I-Associated Mpox Cases Associated with Sexual Contact, the Democratic Republic of the Congo. Emerg Infect Dis. 2024;30(1):172–6.

18. INSP-RDC. Situation report No. 22, Data from epidemiological week 24. 2024.

19. Kinganda-Lusamaki E A-AA, Fernandez N, Makangara-Cigolo JC, Pratt C et al. Clade I Mpox virus genomic diversity in the Democratic Republic of the Congo, 2018 - 2024: Predominance of Zoonotic Transmission. medRxiv. 2024.

20. Migration IU. Democratic Republic of the Congo Crisis Response Plan 2023. International Organization for Migration (IOM); 2023.

21. WHO. Mpox (monkeypox) outbreak toolbox - Updated July 2024. World Health Organization; 2024 July 2024.

22. Minh BQ, Schmidt HA, Chernomor O, Schrempf D, Woodhams MD, von Haeseler A, et al. IQ-TREE 2: New Models and Efficient Methods for Phylogenetic Inference in the Genomic Era. Mol Biol Evol. 2020;37(5):1530–4.

23. Suspene R, Raymond KA, Boutin L, Guillier S, Lemoine F, Ferraris O, et al. APOBEC3F Is a Mutational Driver of the Human Monkeypox Virus Identified in the 2022 Outbreak. J Infect Dis. 2023;228(10):1421–9.

24. O’Toole A, Neher RA, Ndodo N, Borges V, Gannon B, Gomes JP, et al. APOBEC3 deaminase editing in mpox virus as evidence for sustained human transmission since at least 2016. Science. 2023;382(6670):595–600.

25. Masirika LM, Udahemuka JC, Schuele L, Ndishimye P, Otani S, Mbiribindi JB, et al. Ongoing mpox outbreak in Kamituga, South Kivu province, associated with monkeypox virus of a novel Clade I sub-lineage, Democratic Republic of the Congo, 2024. Euro Surveill. 2024;29(11).

26. Shapiro B, Rambaut A, Drummond AJ. Choosing appropriate substitution models for the phylogenetic analysis of protein-coding sequences. Mol Biol Evol. 2006;23(1):7–9.

